# CFTR structures bound to ETI components predict rare mutation response to modulator combinations

**DOI:** 10.1101/2024.10.12.24314979

**Authors:** Noemie Stanleigh, Michal Gur, Michal Shteinberg, Aryeh Weiss, Naama Sebbag-Sznajder, Deborah Duran, Myriam Grunewald, Liron Birimberg-Schwartz, Ronen Bar-Yoseph, Jeffrey M. Beekman, Eitan Kerem, Michael Wilschanski, Batsheva Kerem

## Abstract

**Background:** CFTR protein structures bound to each of Elexacaftor/Tezacaftor/Ivacaftor (ETI) were recently established. We aimed to use this data to predict and assess responses to ETI and each of its components in intestinal organoids derived from patients carrying rare CFTR mutations, not yet approved for ETI, based on their mutation location within the CFTR structure.

**Methods:** Organoids were generated from six patients carrying the Q1100P and/or K163E alleles, not receiving ETI. Measurements of the response to ETI or combination of its components were performed in 3D-organoids by forskolin-induced swelling (FIS) and in 2D-monolayers by short-circuit currents (Isc). Based on these results, patients initiated off-label ETI treatment. Clinical data before and after treatment were collected.

**Results:** VX-445 binds amino acids flanking Q1100P and VX-661 binds near the TM2-ICL1 boundary, where K163E is located. Thus, each modulator was predicted to contribute to the correction of one mutation. Functional measurements (FIS and Isc) indeed showed that Q1100P responded to VX-445 alone, and K163E to VX-661 alone. Unexpectedly, VX-445 had a dramatic effect on K163E function. Both mutations achieved clinically significant CFTR activity levels with VX-661+VX-445, without benefit from VX-770. Following these results ETI was initiated, resulting in significant and sustained clinical improvements, in all patients, in lung function (FEV_1_, LCI), BMI and sweat chloride.

**Conclusion:** These results suggest that our structural approach can help predict response to the available modulators in patients carrying rare CFTR mutations. Furthermore, this approach allows for patient-specific optimization of modulator combinations, minimizing unnecessary exposure to ineffective treatments.

## Introduction

Cystic fibrosis (CF) is a lethal autosomal recessive disease caused by mutations in the cystic fibrosis transmembrane conductance regulator (CFTR) gene, which encodes a chloride ion channel in epithelial cell membranes. The disease is characterized by chronic progressive lung disease, pancreatic insufficiency, gastrointestinal malabsorption, malnutrition, diabetes and male infertility. Over 2100 CFTR sequence variants were identified [1], of which 804 were shown as disease-causing [2].

Substantial progress has been made in the development of CFTR modulating therapies [3]. The latest, highly efficient treatment for mutations affecting CFTR processing and folding, consists of the correctors Elexacaftor (VX-445), and Tezacaftor (VX-661), improving folding, processing and stability of the F508del protein [4–6], and the potentiator Ivacaftor (VX-770), increasing channel opening [7], hereafter referred to as ETI. Clinical trials show ETI significantly improves lung function in CF patients carrying at least one F508del allele [8, 9]. ETI, known as Trikafta in the USA and Kaftrio in Europe, was approved for patients with the F508del mutation, affecting >85% of patients worldwide. The FDA extended approval to 177 additional mutations based on *in-vitro* studies in Fischer Rat Thyroid (FRT) cells [10].

However, ∼10-15% of CF patients carry rare mutations not approved for ETI. While cDNA-based platforms such as FRT cells may help extend available drugs to such subjects, they do not reflect the CFTR native context and might not be a suitable model for all variants, such as splicing and nonsense mutations, dependent on intronic sequences. Ex-vivo models, such as rectal organoids [11], enable measurement of the CFTR modulator response within the individual genetic background.

Intestinal organoids are sensitive to modulator effects and provide a dynamic CFTR-dependent functional readout, measured by the forskolin-induced swelling assay (FIS) [12]. Importantly, *in-vitro* CFTR modulator responses in organoids correlate with lung disease severity and sweat chloride levels, biomarkers of *in-vivo* CFTR function [13, 14]. Recently, structures of the CFTR protein bound to each of the ETI components were established through cryo-electron microscopy, explaining how each modulator rescues the CFTR structure and function [4, 6]. VX-809 and its structural analogue VX-661 stabilize the CFTR by binding to a hydrophobic pocket in TMD1 [6]. VX-445 interacts with the lasso motif and helices TM10 and 11, which interact with NBD1, an interface crucial for protein assembly and the transmission of conformational changes throughout the protein domains [4]. VX-770 binds at a hinge region in TM8, facilitating CFTR channel gating [15]. We used this structural data to predict responses of patients carrying the rare CFTR mutations, Q1100P (c.3299A->C) and K163E (c.487A->G), to ETI treatment and each of its components, based on the location of their mutation within the CFTR structure.

Here we show that both mutations are responsive in intestinal organoids to ETI. As expected from our structural prediction, Q1100P was responsive to VX-445 treatment alone, and K163E responded to VX-661 alone. Unexpectedly, VX-445 alone had a dramatic effect on the function of the K163E allele. Both mutations achieved clinically significant CFTR activity levels with VX-661+VX-445, without benefit from VX-770. This approach allows for patient-specific optimization of modulator combinations, minimizing unnecessary exposure to ineffective treatments. Following these results ETI was initiated, resulting in significant and sustained clinical improvements, in all patients.

## Materials and Methods

### Rectal biopsies and organoid generation

Six patients carrying the Q1100P and/or the K163E alleles were invited to participate in the study (Table 1) and informed consent was obtained. Genotypes were defined by exome sequencing, revealing no additional mutations or polymorphisms within or outside the coding exons (Gene by Gene). Rectal biopsies were obtained at Hadassah Medical Centre and organoids were successfully derived from all patients according to published protocols [12]. Organoids were seeded in Matrigel drops (Corning Inc.), cultured in IntestiCult (Stemcell Technologies) and passaged weekly.

**Table 1:**
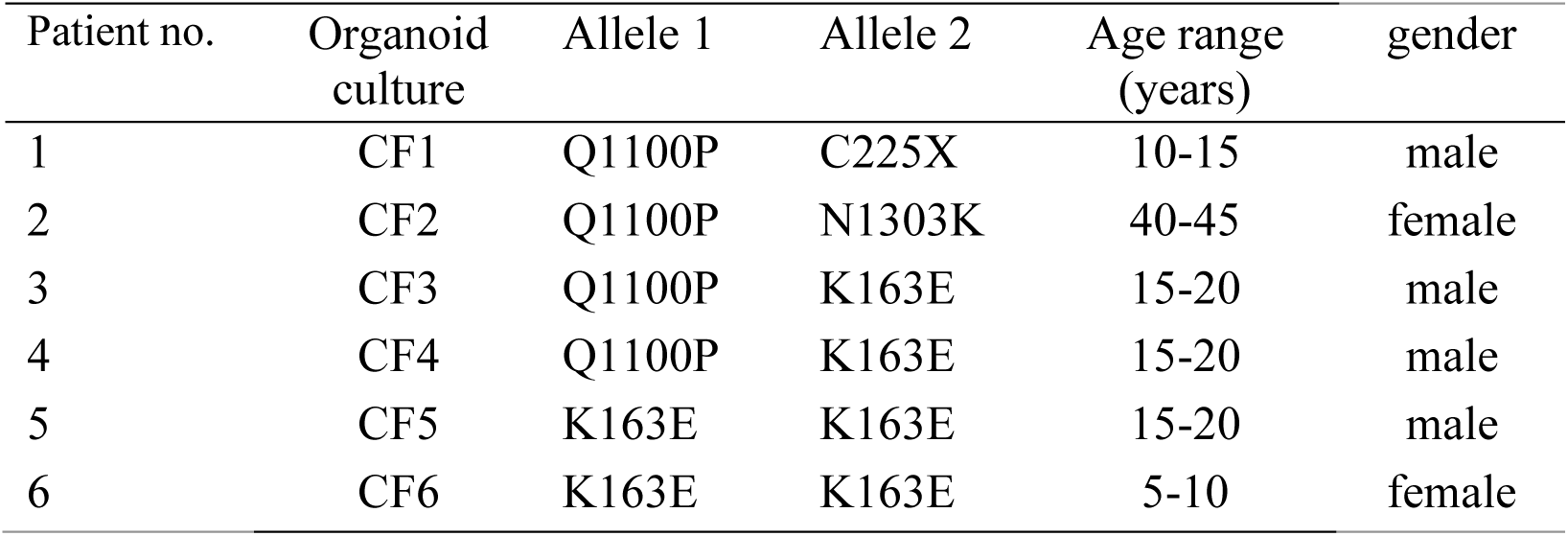
Patient characteristics.

| Patient no. | Organoid culture | Allele 1 | Allele 2 | Age range (years) | gender |
| --- | --- | --- | --- | --- | --- |
| 1 | CF1 | Q1100P | C225X | 10-15 | male |
| 2 | CF2 | Q1100P | N1303K | 40-45 | female |
| 3 | CF3 | Q1100P | K163E | 15-20 | male |
| 4 | CF4 | Q1100P | K163E | 15-20 | male |
| 5 | CF5 | K163E | K163E | 15-20 | male |
| 6 | CF6 | K163E | K163E | 5-10 | female |

### CFTR function measurement in intestinal organoids by FIS

Organoids were incubated with or without 3μM VX-445 and/or VX-661 (Selleck Chemicals LLC) for 24 hours before the assay. FIS was conducted as previously described [12]. The total organoid-area increase was analysed using an in-house Python script in FiJi (see supplement) [16]. The collected data were analysed using Prism10 (GraphPad Software, LLC). Results are presented as mean+SEM of 4-12 biological repeats. Difference between treatments were determined by one-way ANOVA (p<0.05).

### Organoid-derived monolayers and short-circuit current (Isc) recordings

Monolayers were cultured following a published protocol [17] except for our use of Matrigel 1:40 for filter coating. Electrophysiological measurements were performed 6-8 days after seeding, according to SOP ICM_EU001 of the ECFS DNWG, V.2.7 with modifications for monolayers (see supplement). Results are presented as median+min/max range of 4-11 biological repeats, with differences between treatments determined by t-test or one-way ANOVA (p<0.05). Non-CF values were determined in monolayers derived from 4 non-CF individuals who had biopsies taken while undergoing routine colonoscopies. Graphs show non-CF range (grey) and median of the 4 individuals (dotted line), used for calculation.

### CFTR protein levels

Protein extracts were prepared in RIPA buffer separated on 6% poly-acrylamide gels and transferred to a nitrocellulose membrane. Antibody hybridization and chemiluminescence were performed according to standard procedures, using mouse anti-CFTR 596 (CFF) and rabbit anti-calnexin (Sigma) primary antibodies. Secondary antibodies were obtained from Jackson Immunoresearch Laboratories.

## Results

### Genotypes and clinical characteristics of patients participating in the study

We studied the response to the current available modulators in organoids derived from six patients carrying at least one copy of the rare CFTR mutations Q1100P or K163E. Patients 1 and 2 carry Q1100P on one allele and a minimal function mutation on the other allele (Table 1).

Q1100P was reported in Spain, Portugal and Brazil [18–21] and was identified in various population screenings [22–27], although it does not appear in the CFTR2 database. K163E has not been reported in CFTR1 nor CFTR2.

### The triple combination VX-661+VX-445+VX-770 restored CFTR function in organoids with the Q1100P and/or K163E mutations

To assess CFTR function following treatment with the triple combination VX-661+VX-445+VX-770, organoids were incubated with increasing forskolin concentrations (0.02-5μM). No baseline swelling was detected in any of the organoids, indicating minimal function genotypes (Figure 1). Organoids were incubated for 24 hours with 3μM VX-445 and VX-661, and acutely stimulated with forskolin and 3μM VX-770 at time=0. As a control for a clinically relevant response, we used the CFTR activity level in organoids from a patient heterozygous for F508del and the S1251N gating mutation, treated with VX-770 (Figure 1). In order to enable comparison among organoids, we used FIS levels reached at 0.32μM forskolin, which allowed for the best differentiation between responses to treatment. All organoids showed a significant response to VX-661+VX-445+VX-770, ranging from 54-157% of control levels (Table 2). Notably, some functional rescue was observed at the lowest forskolin concentration (0.02µM) in organoids carrying at least one K163E allele, indicating high susceptibility of this mutation to correction (Figure 1).

**Figure 1.**
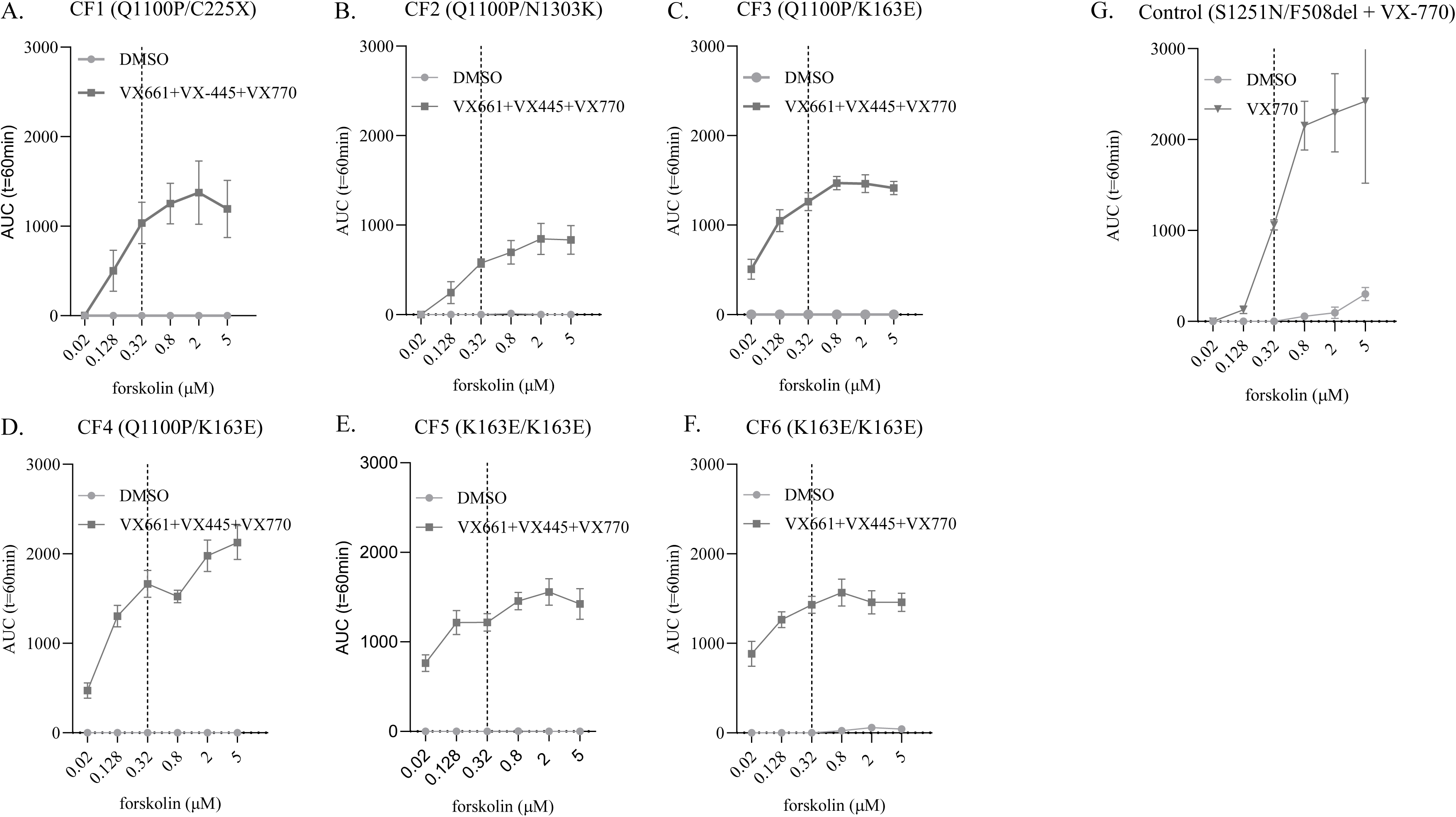
The triple combination VX-661+VX-445+VX-770 restored CFTR function in intestinal organoids carrying the Q1100P and/or K163E mutations. A-F: Forskolin-induced swelling of patient-derived intestinal organoids carrying the Q1100P and/or K163E mutations, treated for 24h with 3µM VX-661+VX-445 or DMSO, induced by different forskolin concentrations, with or without acute addition of 3µM VX-770, expressed as the area under the curve (AUC). G. Control: Forskolin-induced swelling of intestinal organoids derived from a patient compound heterozygous for the S1251N and F508del CFTR mutations, treated with 3µM VX-770. 0.32µM forskolin (dotted line) was chosen for further comparison.

**Table 2:** FIS summary. Results are presented as % of control at 0.32µM forskolin.

| Organoid culture | Basal | VX-661 | VX-445 | VX-661+<br>VX-445 | VX-770 | VX-661<br>+VX-770 | VX-445<br>+VX-770 | VX-661+VX-<br>445+VX-770 |
| --- | --- | --- | --- | --- | --- | --- | --- | --- |
| CF1 | 0 | 0 | 14 | 93 | 0 | 0 | 31 | 98 |
| CF2 | 0 | 0 | 3 | 43 | 0 | 0 | 8 | 54 |
| CF3 | 0 | 12 | 93 | 152 | 0 | 51 | 126 | 119 |
| CF4 | 0 | 21 | 109 | 168 | 1 | 66 | 169 | 157 |
| CF5 | 0 | 10 | 53 | 118 | 1 | 31 | 74 | 115 |
| CF6 | 0 | 50 | 125 | 153 | 7 | 128 | 155 | 135 |

### The double combination VX-661+VX-445 restored CFTR function in organoids with the Q1100P mutation, without added benefit from VX-770

The Q1100P mutation is located in TM11, encoded by exon 20, and substitutes the amino acid glutamine with proline. The CFTR structure in complex with VX-445 shows binding to amino acids 1098 and 1102, flanking Q1100P in TM11 [4]. Thus, VX-445 binding may contribute to the correction of the structural defect introduced by Q1100P, leading to functional correction of the protein. To test our structural predictions, we treated organoids with each modulator separately and in combination.

Patient 1 is a compound heterozygote for the missense mutation Q1100P and the nonsense mutation C225X. C225X introduces a premature termination codon in exon 6 triggering transcript degradation by the nonsense-mediated mRNA decay pathway (NMD) leading to low levels of truncated CFTR proteins. Consequently, C225X cannot contribute to CFTR function. As can be seen in Figure 2, VX-661 alone had no effect on CFTR function in CF1. As predicted from the structural data, VX-445 alone partially restored CFTR function, and addition of VX-770 had no effect. Combined treatment of VX-445 and VX-661 further enhanced CFTR function to 93% of control. Acute addition of VX-770 to the VX-661+VX-445 combination had no beneficial effect (Figure 2, Table 2).

**Figure 2.**
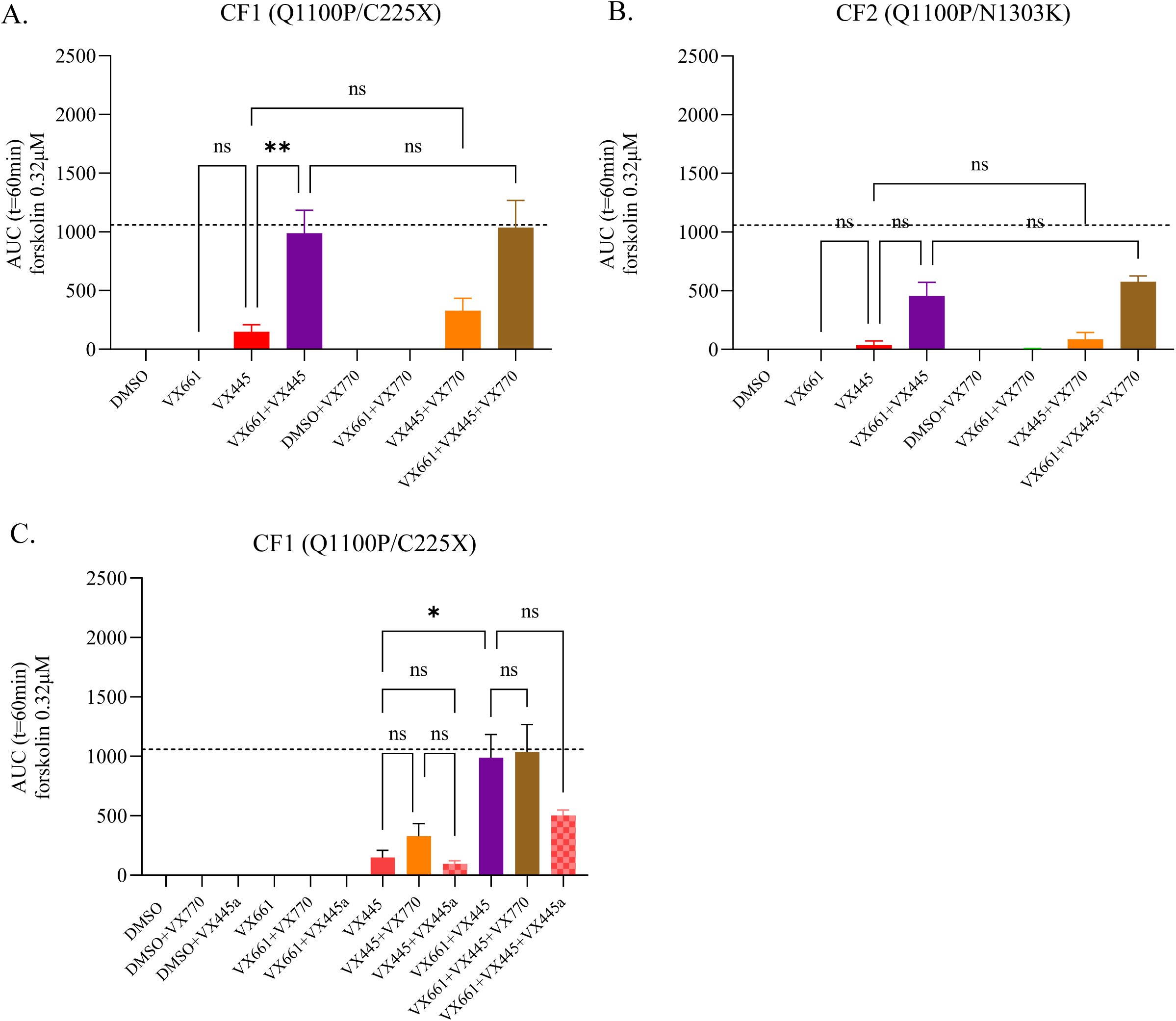
The double combination VX-661+VX-445 restored CFTR function in intestinal organoids carrying the Q1100P mutation, without added benefit from VX-770. A-B: Forskolin-induced swelling of intestinal organoids treated for 24h with 3µM CFTR modulator combinations as indicated, induced by 0.32µM forskolin, with or without acute addition of 3µM VX-770. C. Same experimental conditions as in A-B, except for additional analyses of acute addition of 3µM VX-445 (VX-445a). Data are expressed as mean + SEM of the area under the curve (AUC). Dotted line represents CFTR activity from S1251N/F508del intestinal organoids treated with 3µM VX-770, for a clinically relevant response.

Patient 2 is a compound heterozygote for Q1100P and the minimal function mutation N1303K. N1303K was previously shown by us and others to respond to the triple combination, in patient-derived organoids and primary respiratory epithelial cells [5, 28–31]. The response pattern to modulator treatment in CF2 reflected that of CF1. VX-445 alone had a small effect on CFTR function, which was significantly enhanced with the combined treatment VX-661+VX-445, without additional benefit from VX-770 (Figure 2, Table 2). In comparison, organoids from a patient homozygous for N1303K required all three modulators for restoration of CFTR function (Figure S1), indicating that Q1100P determines the response pattern to modulator treatment in CF2. Maximal CFTR activity in CF2 was moderate, reaching 54% of control.

VX-445 is known to act both as a corrector and a potentiator of WT as well as CFTR proteins with F508del or other rare mutations [30, 32]. To investigate whether the rescue of CFTR function of Q1100P by VX-445 involves potentiation, we examined the effect of acute VX-445 addition on CFTR function in CF1, with and without prior treatment with modulator combinations. Similar to VX-770, no additive effect from acute treatment with VX-445 was seen following treatment with VX-661 alone or in combination with VX-445 (Figure 2). This suggests Q1100P causes a processing/folding defect corrected by VX-661+VX 445. However, the results cannot exclude the possibility that part of the VX-445 effect results from its function as a potentiator of the corrected CFTR proteins.

To evaluate the impact of modulator treatment on mature CFTR protein production, we performed Western blots alongside functional data analysis. The CFTR protein appears as a fully glycosylated band C and incompletely glycosylated band B. Figure 3 shows VX-661 alone did not affect CFTR maturation in CF1. VX-445 enabled band C production, further enhanced by VX-661+VX-445. Weaker signals in CF2 correlated with lower functional rescue. Remarkably, even low CFTR maturation levels led to significant CFTR activity restoration.

**Figure 3.**
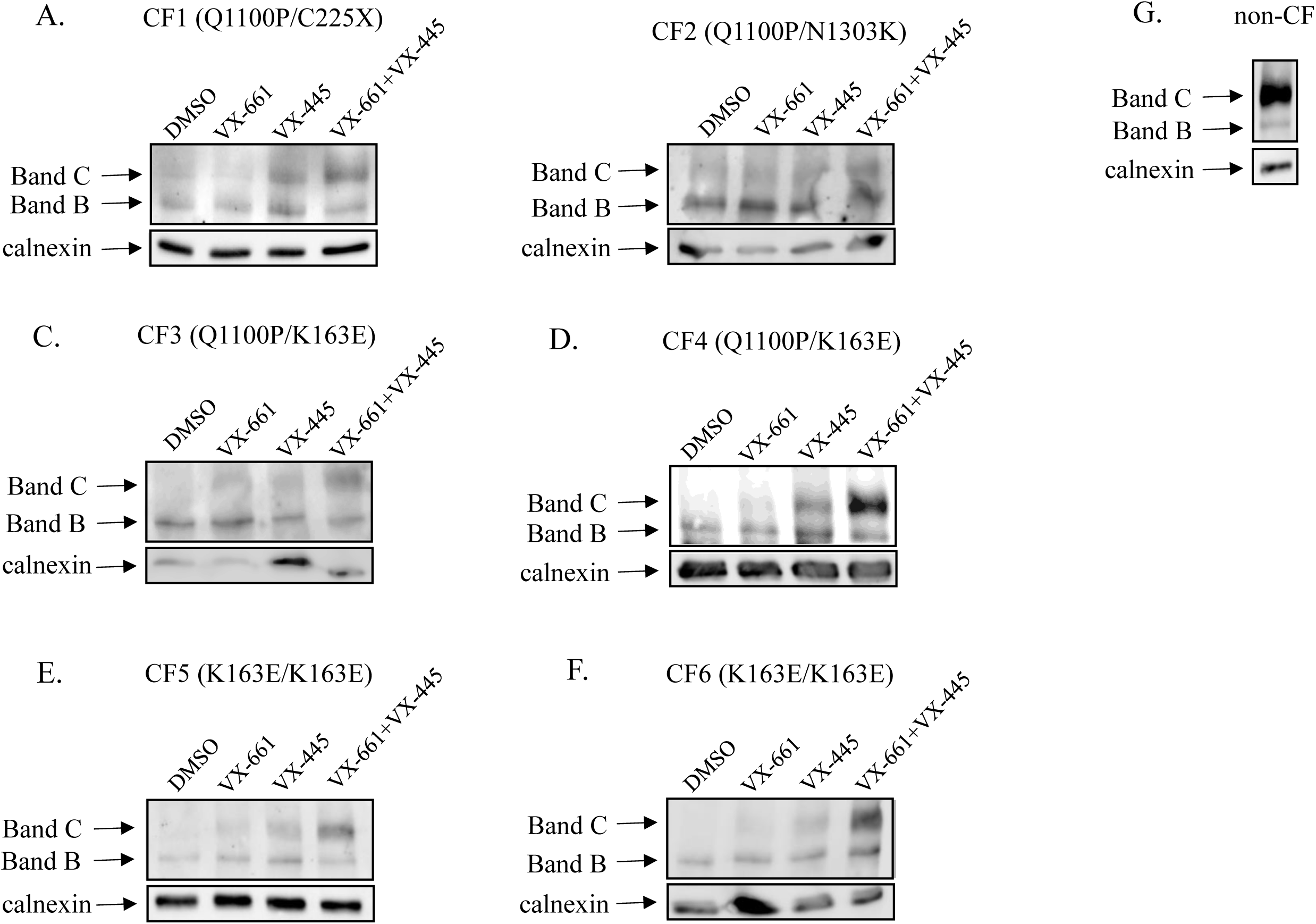
Treatment with VX-661+VX-445 enabled maturation of CFTR proteins in intestinal organoids with the Q1100P and/or K163E mutations. A-F: Western blot analysis of CFTR protein expression in intestinal organoids carrying the Q1100P and/or K163E mutations, after 24-hour treatment with DMSO or 3µM CFTR modulator combinations as indicated. G: Western blot analysis of CFTR protein expression in intestinal organoids derived from a non-CF patient. Calnexin is a loading control.; Band B: immature, core-glycosylated CFTR protein. Band C: mature, complex-glycosylated CFTR protein.

### VX-445 alone significantly restored CFTR function in organoids with the K163E mutation

The K163E mutation is located in exon 4, at the TM2-ICL1 boundary and substitutes lysine by glutamate. The structural data shows that VX-661 binds amino acids in TM2 and TM3 flanking the TM2-ICL1 boundary [6]. Thus, VX-661 binding may contribute to the correction of the structural defect introduced by K163E, leading to functional correction of the protein. CF3 and CF4 are compound heterozygous for the Q1100P/K163E mutations, thus their response may arise from both alleles. CF5 and CF6 are homozygous for K163E. As predicted, VX-661 alone partially restored CFTR function in organoids from all patients (CF3-CF6) and acute VX-770 addition enhanced this CFTR activity (Figure 4, Table 2). Surprisingly, VX-445 alone significantly restored CFTR function reaching control levels in CF3, CF4 and CF6. The response of CF6 reached its maximum effect already with VX-445 alone. The addition of VX-661 further enhanced the response to VX-445 in CF3-CF5, exceeding control levels. In all organoids, VX-770 addition provided no additional benefit. This is a different response pattern than the one observed for Q1100P, which did not respond to VX-661 alone (Figure 2). These results suggest that K163E determines the response pattern in all cultures carrying this mutation.

**Figure 4.**
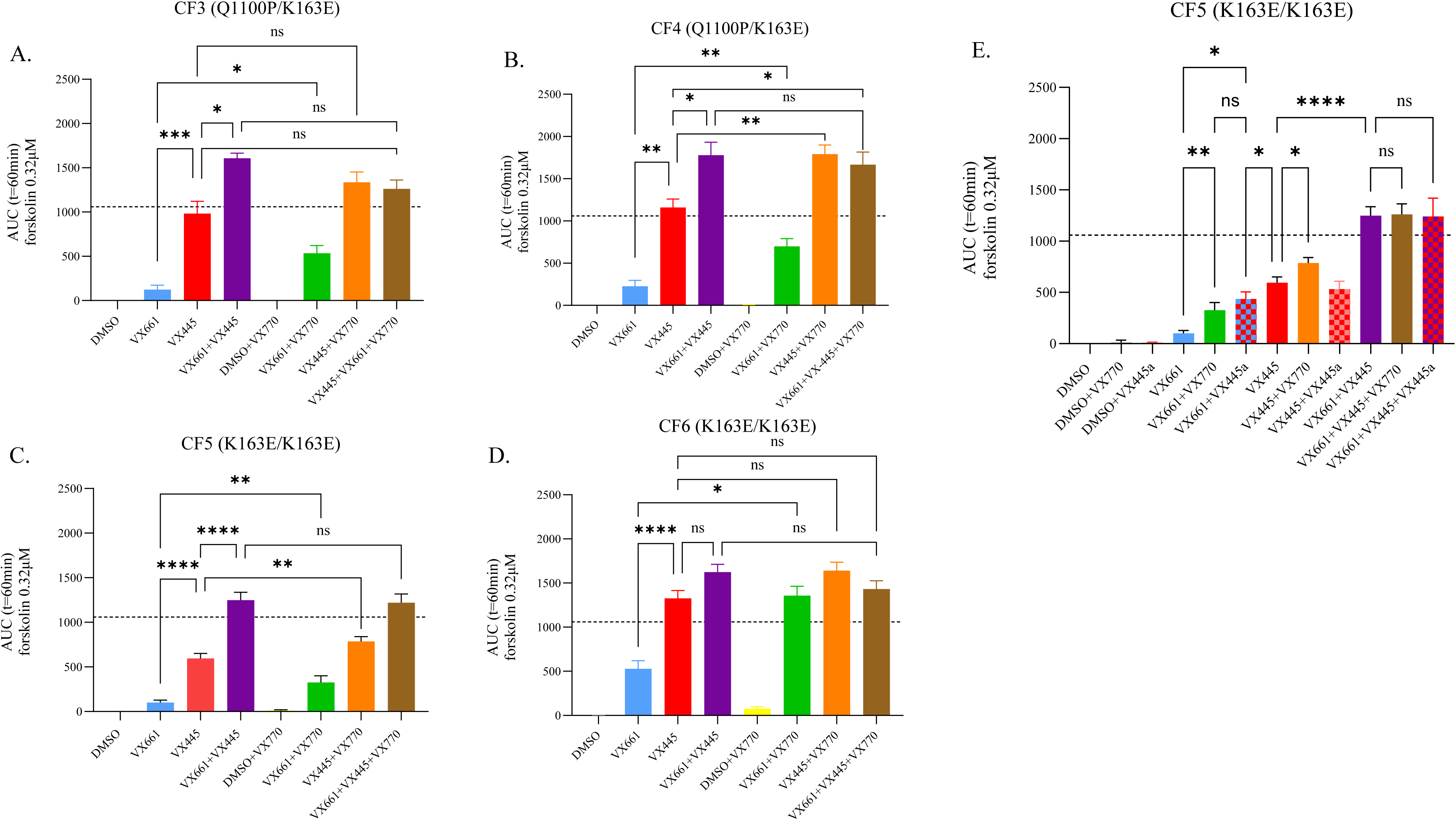
VX-445 alone significantly restored CFTR function in intestinal organoids with the K163E mutation. A-D: Forskolin-induced swelling of intestinal organoids treated for 24h with 3µM CFTR modulator combinations as indicated, and induced by 0.32µM forskolin, with or without acute addition of 3µM VX-770. E. Same experimental conditions as in A-D, except for additional analyses of acute addition of 3µM VX-445 (VX-445a). Data are expressed as mean + SEM of the area under the curve (AUC). Dotted line represents CFTR activity from S1251N/F508del intestinal organoids treated with 3µM VX-770, for a clinically relevant response.

To investigate whether the rescue of CFTR function of K163E by VX-445 involves potentiation, we examined the effect of acute addition of VX-445 on CFTR function on K163E. The analysis was performed in CF5, homozygous for the mutation. Acute VX-445 addition significantly increased CFTR activity after chronic VX-661 treatment (Figure 4), indicating VX-445 acted as a potentiator of the VX-661-corrected K163E protein. However, this potentiation resulted in a lower activity than chronic treatment with VX-445 alone. Altogether, each corrector alone benefited from the addition of a potentiator, but the highest CFTR activity was achieved by the combination of the two correctors VX-661+VX-445, suggesting K163E causes a processing/folding defect. However, we cannot exclude the possibility that part of the chronic VX-445 effect results from its function as a potentiator of the corrected CFTR proteins.

Analysing the effect of modulator treatment on CFTR protein maturation by Western blots of CF3-CF6 showed VX-661 alone partially produced mature CFTR proteins, and VX-445 alone had a stronger effect, in all organoids, but to a lesser extent in CF3. This confirms that VX-445 effectively corrected the processing defect induced by K163E. VX-661+VX-445 further increased CFTR maturation (Figure 3). Despite variations in CFTR maturation levels, CFTR function in all organoids reached or exceeded control levels, indicating that significant function restoration can be achieved even by partial maturation.

### The double combination VX-661+VX-445 restored CFTR function in intestinal monolayers, without added benefit from VX-770

Our results using FIS show a restoration of CFTR activity similar or higher than the response of gating organoids treated with VX-770, suggesting a clinical relevance. For further validation of the response pattern of Q1100P and K163E, we analysed CFTR response to the modulators by Isc, using 2D-cultures from the same organoids, enabling a direct measurement of CFTR function as well as comparison of the response to non-CF levels. Measurements in CF1 (Q1100P/C225X) monolayers showed no response to VX-661, partial response to VX-445 and the response was further enhanced by VX-661+VX-445, reaching ∼60% of the non-CF response (Figure 5). This response pattern was similar to that observed in FIS. Measurements in CF4 (Q1100P/K163E) and CF5 (K163E/K163E) monolayers showed partial response to VX-661, significant response to VX-445 alone, reaching 79% and 84% of non-CF level respectively, and the response was further enhanced by VX-661+VX-445, reaching above non-CF range (106% and 118% of non-CF) (Figure 5). In these monolayers, acute VX-770 addition had no beneficial effect. This response pattern was similar to that observed in FIS. It is important to note that there is a high correlation between the responses in FIS and Isc to each treatment (Figure S2), indicating that Q1100P does not benefit from VX-770 addition and K163E is primarily responding to VX-445. Furthermore, the Isc measurements, which reached non-CF levels, support the clinical potential of modulator treatment.

**Figure 5.**
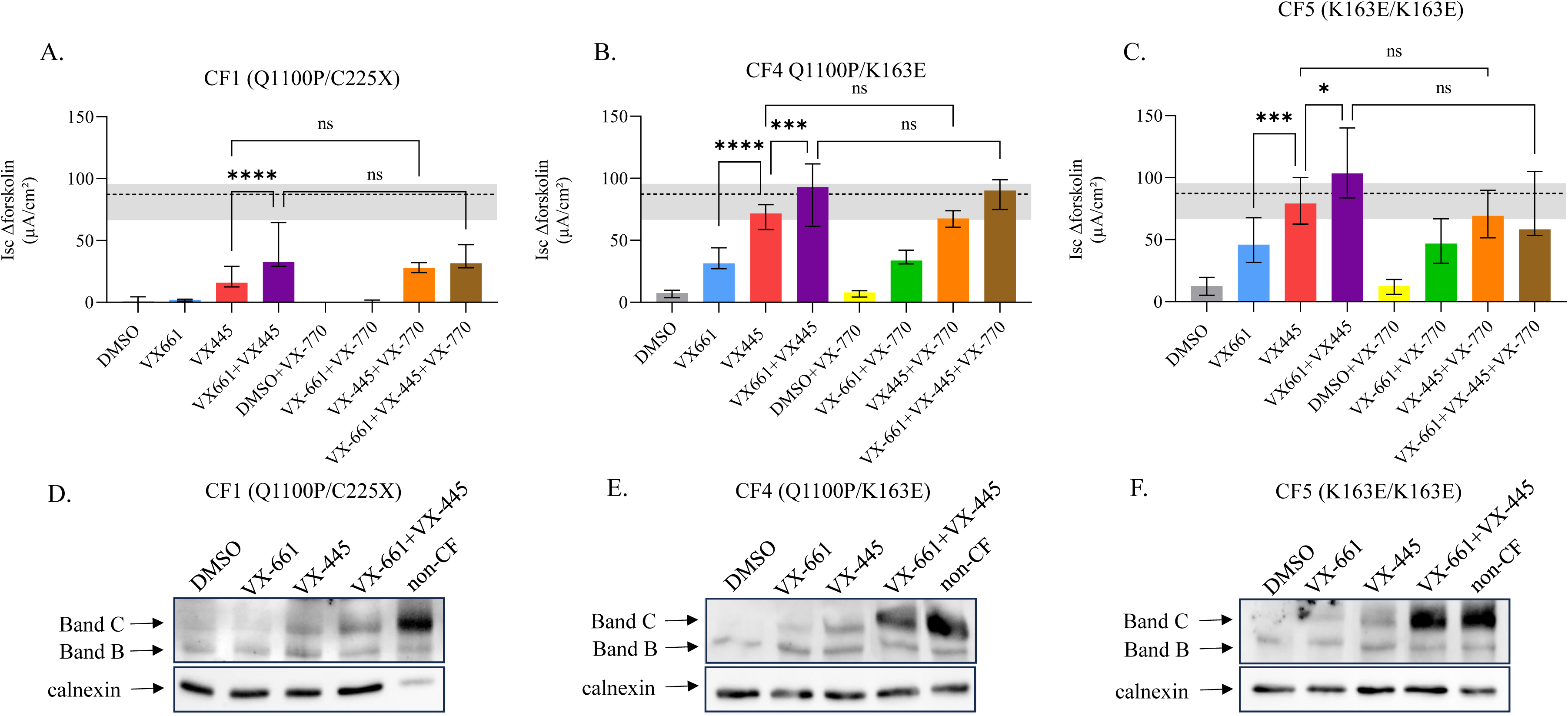
The double combination VX-661+VX-445 restored CFTR function in intestinal monolayers carrying the Q1100P and/or K163E mutations, without added benefit from VX-770. A-C: Short-circuit currents (Isc) of intestinal organoid monolayers treated for 24h with 3µM CFTR modulator combinations as indicated and induced by 10µM forskolin, with or without acute addition of 3µM VX-770. Results are presented as the median + min-max range of Δforskolin. In grey: non-CF range, dotted line: non-CF median. D-F: Western blot analysis of CFTR protein expression in intestinal organoid monolayers after 24-hour treatment with 3µM CFTR modulator combinations as indicated, or DMSO. Calnexin is a loading control. Band B: immature, core-glycosylated CFTR. Band C: mature, complex-glycosylated CFTR.

Analysis of CFTR protein maturation in CF1 monolayers showed VX-661 alone did not affect CFTR maturation, VX-445 increased band C production, further enhanced by VX-661+VX-445, similarly to the pattern observed in 3D organoids (Figure 5). Analysis of CFTR protein maturation in CF4 an CF5 showed VX-661 alone induced CFTR maturation, VX-445 further increased band C production, and treatment with VX-661+VX-445 restored protein levels to non-CF levels. The higher level of protein maturation observed after modulator treatment in monolayers compared to 3D organoids might be explained by the exposure to the medium of both apical and basal side of cells in monolayers, compared to the 3D medium exposure from the basal side only, resulting in higher modulator uptake and efficiency. Altogether, the CFTR response pattern measured in monolayer organoids validated the pattern found in the 3D model.

### ETI treatment had a significant clinical benefit in patients with the Q1100P and/or the K163E mutations

All patients showed significant clinical improvement after one month of treatment with ETI: a 24.2%±14.7 mean increase in FEV_1_, a 2±2.5kg mean increase in weight, a 0.8±0.4 mean increase in BMI among the 5 adult patients (1-5), a −4.0±1.6-mean decrease in LCI, and a −23.3mmol/L±13.1 mean decrease in sweat chloride (Table 3A). In four out of five patients, sweat chloride levels were reduced to normal or borderline levels. All patients had long-term follow-up (at two time points) in pulmonary function, BMI and weight, showing sustained improvement in FEV_1_ and continued increase in BMI and weight (Table 3B).

**Table 3:** Clinical data before and after 1 month of treatment with ETI. A. Effect of ETI treatment after one month. B. Long term effect of ETI treatment.

|  | PS/PI | CFRD | FEV<sub>1</sub>, % predicted |  |  | Sweat chloride, mmol/l |  |  |
| --- | --- | --- | --- | --- | --- | --- | --- | --- |
| Patient no. |  |  | Before | After 1 month | Change | Before | After 1 month | Change |
| 1 | PI | - | 79 | 113 | +34 | 102 | 86 | -16 |
| 2 | PI | - | 53 | 99 | +46 |  |  |  |
| 3 | PS | - | 53 | 74 | +21 | 72 | 42 | -30 |
| 4 | PS | - | 75 | 90 | +15 | 73 | 64 | -9 |
| 5 | PS | - | 76 | 101 | +25 | 48 | 22 | -26 |
| 6 | PS | - | 99 | 103 | +4 | 70 | 32 | -38 |
| average |  |  | +24.2±14.7 |  |  | -23.8±11.5 |  |  |

|  | LCI |  |  | BMI |  |  | Weight (kg) |  |  |
| --- | --- | --- | --- | --- | --- | --- | --- | --- | --- |
| Patient no. | Before | After 1 month | Change | Before | After 1 month | Change | Before | After 1 month | Change |
| 1 | 10.7 | 7.5 | -3.2 | 19.7 | 20.3 | +0.6 | 60.2 | 62.5 | +2.3 |
| 2 |  |  |  | 19 | 20 | +1 | 54.2 | 56.9 | +2.7 |
| 3 | 16.4 | 11.5 | -4.9 | 20.9 | 22.2 | +1.3 | 61.8 | 68 | +6.2 |
| 4 | 13.1 | 7.5 | -5.6 | 27.1 | 28.2 | +1.1 | 87 | 87.3 | +0.3 |
| 5 | 13.7 | 9.7 | -4 | 25 | 24.9 | -0.1 | 76.6 | 75.5 | -1.1 |
| 6 | 8.3 | 6.1 | -2.2 | 16.6 | 16.8 | +0.2 | 28 | 29.8 | +1.8 |
| average | -4±1.3 |  |  | 0.7±0.5 |  |  | 2±2.5 |  |  |

Abbreviations: PS - pancreatic sufficient; PI - pancreatic insufficient; CFRD – CF-related diabetes, FEV<sub>1</sub>- forced expiratory volume in 1 second, LCI – lung clearance index, BMI – body mass index.

B. Long term effect of ETI treatment
| Patient no. | FEV<sub>1</sub>, % predicted |  |  |  |
| --- | --- | --- | --- | --- |
|  | Before | After*<br>1<sup>st</sup> follow-up (5-19m) |  | After*<br>2<sup>nd</sup> follow-up ( |
| 1 | 79 | 104 (9m) | +25 | 105 (15m) +26 |
| 2 | 53 | 97 (5m) | +44 | 96 (14m) +43 |
| 3 | 53 | 69 (19m) | +16 | 71 (26m) +18 |
| 4 | 75 | 88 (19m) | +13 | 88 (23m) +13 |
| 5 | 76 | 94 (15m) | +18 | 102 (18m) +26 |
| 6 | 99 | 104 (8m) | +5 | 97 (11m) -2 |

BMI
| Patient no. | BMI |  |  |  |
| --- | --- | --- | --- | --- |
|  | Before | After*<br>1<sup>st</sup> follow-up |  | After*<br>2<sup>nd</sup> follow-up |
| 1 | 19.7 | 21.2 (9m) | +1.5 | 21.8 (15m) +2.1 |
| 2 | 19 | 21 (5m) | +2 | - |
| 3 | 20.9 | 26.1 (19m) | +5.2 | 25.4 (26m) +4.5 |
| 4 | 27.1 | 29.4 (19m) | +2.3 | 31.2 (23m) +4.1 |
| 5 | 25 | 25.6 (15m) | +0.6 | 25.8 (18m) +0.8 |
| 6 | 16.6 | 15.9 (8m) | -0.7 | 16.9 (11m) +0.3 |

| Patient no. | Weight (kg) |  |  |  |
| --- | --- | --- | --- | --- |
|  | Before | After*<br>1<sup>st</sup> follow-up |  | After*<br>2<sup>nd</sup> follow-up |
| 1 | 60.2 | 65 (9m) | +4.8 | 68.3 (15m) +8.1 |
| 2 | 54.2 | 59 (5m) | +4.8 | - |
| 3 | 61.8 | 80 (19m) | +18.2 | 78.6 (26m) +16.8 |
| 4 | 87 | 92 (19m) | +5 | 97.8 (23m) +10.8 |
| 5 | 76.6 | 78.3 (15m) | +1.7 | 80 (18m) +3.4 |
| 6 | 28 | 29.4 (8m) | +1.4 | 32.7 (11m) +4.7 |

\* In parentheses is the number of months since the beginning of ETI treatment.
LCI and sweat chloride levels were not measured at follow-up appointments until the time of publishing

## Discussion

Here we demonstrated the ability of VX-661, VX-445 and VX-770 to restore CFTR activity in organoids derived from patients carrying the ultra-rare mutations Q1100P and K163E. These mutations were selected based on the CFTR structure and modulator binding sites [4, 6] as a proof of concept for a structure-based approach.

Q1100P, located in TM11, disrupts the α-helix structure due to the bulky side chain of proline and its inability to form hydrogen bonds. This might prevent correct CFTR folding and processing [20]. VX-445 stabilizes TM helices 10 and 11 [4], strengthening the TMD1/NBD1 interface, crucial for CFTR folding and processing [33]. VX-445 interacts with TM11 through residues adjacent to Q1100, potentially correcting the Q1100P defect. Notably, ETI has been approved for the adjacent M1101K mutation based on FRT cells results [10]. Interestingly,VX-445 alone increased M1101K function, which was further augmented by VX-661 [5]. The same response pattern was found in CF1 and CF2, carrying Q1100P (Figure 2). The additional benefit from VX-661 may result from its stabilizing effect of TMD1 during CFTR folding, acting synergistically with VX-445 [5, 6].

K163 is located in TMD1, and is part of a stretch of conserved lysine residues at the TM2-ICL1 boundary [34], a region participating in the ICL1-NBD1 interphase, which is essential for correct folding of the CFTR protein. This region is stabilized by VX-809 [33, 35, 36] and its structural analogue VX-661 [6]. Accordingly, VX-661 partially restored CFTR activity in all organoids carrying K163E (CF3-6, Figure 4). Unpredictably, VX-445 alone achieved a high restoration of CFTR activity in all K163E organoids, suggesting that beside the binding of VX-445 to TM2 at residue I132 it may have additional effects in the domain in which K163E resides [35]. Maximal levels of restoration in K163E organoids were achieved by VX-661+VX-445, suggesting each of the modulators stabilizes the K163E protein by different mechanisms, acting synergistically. Our results may help elucidate the structural basis of modulator responses and assist in understanding regions important for CFTR domain interactions and function.

We assessed CFTR function in patient-derived intestinal epithelial cells using two assays, FIS and Isc. FIS is a relatively simple assay, measuring CFTR function indirectly by coupling ion secretion to fluid transport. Isc in monolayers enable for a direct measurement of CFTR activity and a direct comparison to non-CF activity but require complex culture and measurement methods. We show a significant correlation between the CFTR responses to the different modulator combinations in the two assays (Figure S2), further validating FIS as a reliable assay for CFTR activity. Moreover, the unique response pattern of each of the rare, studied CFTR mutations was observed in both assays, confirming Q1100P and K163E do not benefit from VX-770 addition and K163E is primarily responding to VX-445.

Based on our *in-vitro* results, the medical providers approved off-label treatment with ETI for all patients presented here. The clinical benefits were immediate and substantial. After the first month of treatment, the mean FEV_1_ increased by 24.2%+14.7 (Table 3A). Patients had long-term follow-up in pulmonary function and BMI, showing sustained improvement (Table 3B). Thus, our results further support that results from organoids can serve as a basis for drug approval for patients carrying rare CFTR mutations not included in clinical trials.

In a large study, Bilher et al. analysed the response of 655 rare CFTR variants to ETI, using transient transfections of CFTR variants into FRT cells [37]. In this study, the response of the Q1100P allele was assessed and surprisingly it was defined as non-responder. The threshold defined in that large study was 10% of WT, however, as the authors mentioned, this threshold should not be considered as a rigid cut-off. We suggest that borderline responses should be assessed in patient-derived systems or if possible, get a clinical assessment of ETI responsiveness.

Finally, the highly effective treatments with the current available modulators have side effects. VX-770 has been shown to cause headaches, rash, elevation in liver enzymes [38] and some patients treated with ETI experienced several side effects [8, 9], including serious psychiatric side effects in ∼10% of patients after prolonged treatment [39]. The results presented in this study, as well as previous results studying other rare mutations [40] suggest that the use of patient-derived models can help optimize modulator combinations for rare mutants, avoiding unnecessary patient exposure to ineffective and potentially harmful compounds. Altogether, our results suggest that using data of the CFTR structure and modulator binding sites can predict response to treatment, thus giving patients with rare mutations an opportunity to benefit from existing treatments that have not been specifically approved for their mutation.

## Data Availability

All data produced in the present study are available upon reasonable request to the authors

## Acknowledgements

We are grateful to all CF patients for their participation in the research.

We thank Naomi Melamed-Book from the Bioimaging Unit of the Alexander Silberman Institute of Life Sciences, The Hebrew University of Jerusalem, for the imaging of intestinal organoids. We also thank the entire past and present members of the Kerem group, and especially Michal Tur-Sinai, for technical advice, assistance and enlightening discussions. This study utilized OpenAI’s GPT-4 for assistance in shortening the manuscript.

## Funding

This research was supported (in part) by grants from the Cystic Fibrosis Foundation to

B.K. (KEREM19KO) and M.W. (WILSCH20K0).

## Declaration

Conflict of interests: The authors have no relevant financial or non-financial interests to disclose.

## Notes

### Competing Interest Statement

The authors have declared no competing interest.

### Author Declarations

The ethics committee of the Hadassah University Hospital in Jerusalem, Israel, gave ethical approval for this work.

