## supplement for "CFTR structures bound to ETI components predict rare mutation response to modulator combinations"

**Automated analysis of organoid swelling**

Organoid swelling was measured using a python program that was developed in-house withing the Fiji/ImageJ [16] image processing environment. The program makes use of minimal use input to generate consistent results across different data sets. Important features of the program are:

1. Automated detection of the mean and standard deviation of the background. The threshold for object detection is calculated as the background mean plus a user determined number of standard deviations above background. This method is applied consistently across the entire dataset, so that the user does not subjectively select the segmentation threshold.
2. The program prevents distinct organoids from merging into each other as they swell. Therefore, in addition to mean organoid swelling, the swelling of individual organoids is available in the output.
3. The only other user-selected segmentation parameters are number of erosions (which removed small punctate artifacts) and minimum circularity (which removes line artifacts).
4. Swelling organoids may break up near the end of the time series. These organoids are removed, as are objects that do not appear in all frames of the time series.
5. The program outputs absolute and normalized swelling data for each detected organoid, as well as mean swelling (weighted for organoid area) as a set of CSV files. In addition, an annotated image sequence is saved, to enable visual assessment of the output quality.
   Log files containing the parameters with which the code was run are also saved, as are various intermediate files that assist in debugging, and verification of the results.

It is important to note that the code analyzes two-dimensional images, but the organoids obviously swell in three dimensions. If the culture is too dense, the organoids will swell against each other, at which point no further swelling will be observed. However, the code will prevent them from merging.

The code is available from Aryeh Weiss. It must be noted that the code is currently specific to the file format and directory structure used in this work. Furthermore, the code will be provided as-is, with no guarantee that it will work for the recipient.

**Short-circuit current (Isc) recordings in monolayers**

Filters were treated 24 hours with 3µM VX-661 and/or 3µM VX-445. Filter inserts were mounted in P2302T sliders. The transepithelial potential difference was clamped with a VVC-MC6S module (Physiologic Instruments), and Isc was recorded using Acquire&Analyze (version 2.3), while adding compounds to the basal (B) or apical (A) bathing solutions: 100µM amiloride (A+B), 100µM IBMX and 10µM forskolin (A+B), 0.3µM VX-770 (A), 10µM Inh172 (A+B).

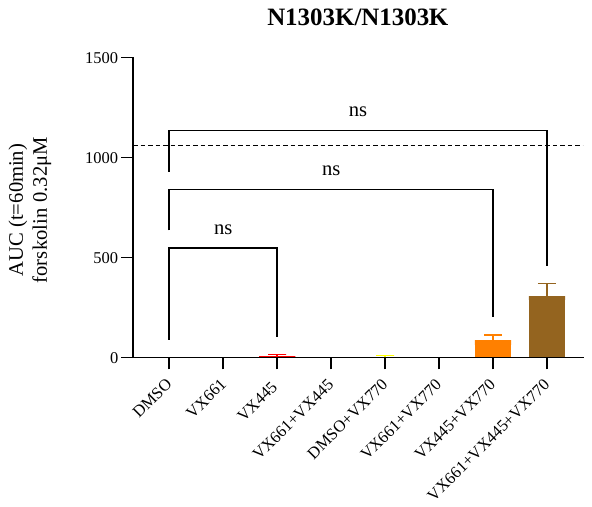

**Figure S1. Restoration of CFTR activity in intestinal organoids from a patient homozygous for the N1303K mutation required the triple combination of VX-661+VX-445+VX-770**

Forskolin-induced swelling of intestinal organoids derived from a patient homozygous for the N1303K mutation treated by 3µM modulator combinations as indicated and induced by 0.32µM forskolin with or without acute addition of 3µM VX-770. Data are expressed as mean + SEM of the area under the curve (AUC). Dotted line represents CFTR activity from S1251N/F508del intestinal organoids treated with 3µM VX-770, as a control for a clinically relevant response.

**Figure S2. Correlation between CFTR activity after modulator treatments as measured by FIS and Isc**

Pearson correlation of intestinal organoid swelling as measured by FIS versus short-circuit current measurements in organoid monolayers (Isc), after 24h treatment with 3µM CFTR modulator combinations and induction with forskolin, with or without acute addition of 3µM VX-770. The analysis was performed in CF1, CF4 and CF5.

Blue – VX-661, Red – VX-445, purple: VX-661+VX-445, Green – VX-661+VX-770, Orange VX-445+VX-770, Brown – VX-661+VX-445+VX-770.

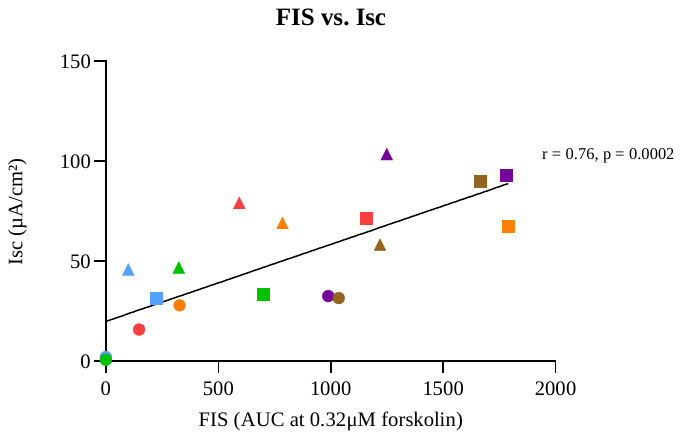

CF1

CF4

CF5
